# Characterizing the relative strength of metapopulation dynamics in measles persistence

**DOI:** 10.64898/2026.10.24.26366357

**Authors:** Deepit Bhatia, Amalie McKee, Matthew Ferrari

**Affiliations:** Pennsylvania State University; Harrisburg University

## Abstract

**Background/Aims:** Measles is a highly infectious, vaccine-preventable disease that remains persistent across much of the world despite continued large-scale control efforts. Local dynamics of measles are shaped by births of susceptible individuals and seasonal drivers of transmission, while broader regional dynamics are shaped by population mobility and metapopulation connectivity. Incidence data, useful for tracking the progress of measles control, are frequently reported only at the aggregate level (e.g., states/provinces), and persistence at this scale may arise because of the independent persistence in all local sub-units or because of regional metapopulation dynamics. Disentangling the contribution of local versus regional persistence is critical in identifying effective strategies for control.

**Methods/Results:** We show, as vaccination reduces local measles prevalence, metapopulation colonization and extinction events dominate regional persistence. This manifests as a change in the seasonal distribution of cases throughout the year; incidence becomes more uniformly distributed through the year even with no change in underlying seasonal drivers. We demonstrate this using: 1) a seasonally forced, stochastic metapopulation compartmental model; 2) historical measles incidence in the USA; and 3) recent subnational-level measles incidence in Nigeria.

We create a novel, scale-free measure to quantify the contribution of metapopulation dynamics to persistence, roundness, defined as the geometric mean of the scaled maximum and minimum monthly incidence. A low value of roundness indicates strong local persistence dominated by the seasonal attractor, while a high value indicates strong metapopulation persistence. We illustrate the properties of this measure using simulations and observed data. In the USA, roundness nearly doubled after the introduction of the measles-containing vaccine. In Nigeria, roundness is well correlated with observed vaccine coverage, with a Pearson’s *r* of 0.70 (95% CI: 0.57, 0.80), discriminating between northern states dominated by local persistence of measles and southern states where persistence is maintained by regional dynamics.

**Conclusion:** Identifying how measles persists is important in its control—vaccination strategies may differ if persistence is largely local, or if persistence is driven by connectivity from surrounding areas, even where yearly incidence is comparable. Our approach can be valuable areas where other measures (connectivity, mobility, laboratory confirmed illness) are not widely available.

## Introduction

Vaccination has drastically reduced the incidence and burden of communicable disease in humans (Klepac et al., 2013). Since 1974, 154 million deaths have been averted, of which two-thirds were in children less than 1 year of age. Vaccination has accounted for 40% of the observed decline in infant mortality worldwide (Shattock et al., 2024). For example, vaccination has averted 1.6 million deaths due to polio, 279 million deaths due to tetanus, and 94 million deaths due to measles (Shattock et al., 2024). Between 1980 and 2009, coverage for routine childhood vaccines including diphtheria/pertussis/tetanus (DTP3), polio, and measles containing vaccine (MCV) increased by 2 percentage points per year (Baidya et al., 2025). However, since the start of the COVID-19 pandemic, progress has slowed and even reversed in some circumstances, with DTP3 coverage now at its lowest level since 2009, and drops in MCV coverage led to a 30-fold increase in measles cases in Europe compared to 2016 (Gambrell et al., 2022; Mahase, 2025). Even as vaccination has increased, there continue to be large gaps both between and within countries (Bhatia et al., 2025). Local heterogeneity in the density of susceptible individuals, due to in variation in vaccination coverage, can increase the vaccination coverage necessary to maintain elimination (Truelove et al., 2019). Movement between and among groups with varying levels of coverage can lead to outbreaks in areas that may otherwise have a high aggregate level of coverage (Keeling, 2000). Addressing these heterogeneities and gaps in coverage, particularly at the subnational and subgroup level, has been an emphasis of the Immunization Agenda 2030 (IA2030) set out by World Health Organization (WHO) (IA2030, 2020).

Incidence data are frequently recorded and reported at the aggregate, or metapopulation level (i.e., administrative units), and persistence may arise either because subunits below the level of reporting are independently persistent, or because of extinction-colonization processes (Grenfell & Harwood, 1997). As available data become more aggregated, disease dynamics at that aggregated, or metapopulation level may not be informative about transmission dynamics at the patch-level within (Gunning & Wearing, 2013; Metcalf et al., 2013). A key dynamical consequence is that even if some local populations, or patches, have an effective reproductive number (R_E_) of less than 1, disease may persist at the regional level due to the extinction-colonization dynamics consistent with a metapopulation (Truelove et al., 2019).

These dynamics are complicated by issues of measurement scale. While efforts have been ongoing to emphasize subnational and subgroup-level gaps in disease control (IA2030, 2020), the scale of disease and coverage reporting can often be mismatched with the scale of disease transmission itself. At smaller spatial scales, the reduced size of the population compounded with low reporting rates can make estimates of persistence and risk inaccurate (Gunning & Wearing, 2013; Olsen et al., 1996). Such measurement issues create distinct control challenges. Gunning et al highlighted that underreporting can make the determination of local persistence difficult, where small to medium sized populations may maintain persistent transmission but present as 0 reported cases in administrative time series due to under-reporting (Gunning et al., 2017). Another challenge is that spatial units at which data are collected and reported are often political, so they may exclude relevant information about disease dynamics and control from neighboring jurisdictions. Therefore, understanding disease dynamics at smaller spatial scales— closer to the scale at which transmission is occurring—is important in assessing control strategies and progress toward disease elimination.

Measles is a highly infectious viral disease of humans caused by the measles virus, causing over 10 million cases and over 100,000 deaths in 2023 alone (Guerra et al., 2017; Minta, 2024; Tesini, 2023). The basic reproductive ratio, R_0_, which quantifies the expected number of secondary infections due to the first infection in a completely susceptible population, of measles is 12-18, higher than many other directly communicable diseases in humans (Guerra et al., 2017). The strong and lasting immunity due to measles infection, coupled with seasonal forcing of transmission due to human behavioral cycles drives cyclic dynamics of outbreaks and frequent local fade-out and metapopulation rescue, even in the pre-vaccine era (Ferrari et al., 2010; Grenfell & Harwood, 1997). The high R_0_ of measles necessitates a high level of vaccination coverage to control, which can be difficult to achieve and maintain uniformly. The reductions in incidence since the introduction of vaccination result in correspondingly lower incidence in the inter-epidemic troughs. Thus, metapopulation dynamics are critical to the regional persistence of measles (Anderson & May, 2010; Bartlett, 1956; Conlan et al., 2010; Earn et al., 2000). Ferrari et al, 2008, showed that metapopulation dynamics can generate regular, persistent cyclic dynamics at the regional level in settings where the annual average R_E_ is greater than 1, but strong seasonal forcing drives low prevalence and high rates of stochastic extinction during periods of seasonal low transmission (Ferrari et al., 2010). While estimates for the critical community size (CCS), or the minimum population size of a community to allow for sustained transmission of measles, exist, it is difficult to quantify how this interacts with seasonality in transmission, metapopulations, and the effect of vaccination (Ferrari et al., 2013; Grenfell & Harwood, 1997). However, in patchy environments, Ferrari et. al have shown that frequent extinctions of measles can occur even in areas with low coverage (Ferrari et al., 2008, 2010).

For seasonally forced pathogens, like measles, we see highly synchronized outbreaks in connected spatial regions (Rohani et al., 1999). This synchrony begins to decline after the introduction of the vaccine, even when the seasonality in transmission remains constant (Rohani et al., 1999). As synchrony declines, even when seasonal forcing is unchanged, the aggregate pattern of incidence over many patches should broaden due to extinction and colonization dynamics (**Figure 1**). At low coverage, outbreaks are likely to be spatially synchronized at the patch level, and with a sufficient supply of susceptible individuals, infection persists through troughs of low seasonal transmission (see **Figure 1**, A). If susceptibles are limited, e.g., through vaccination, then the likelihood of local extinction increases during seasonal troughs. As susceptibles build up over time, the start of the following outbreak will depend both on the seasonal transmission rate as well as the rate of re-introduction. This can give rise to later-than-expected outbreaks if introduction occurs later than the seasonal peak in transmission or earlier than expected if susceptible accumulation leads to an increase in R_E_ above 1. Thus, observing this phenomenon at the metapopulation level, we may see, for the same underlying seasonal transmission, an epidemic curve that is much flatter with respect to seasonal variation.

**Figure 1:**
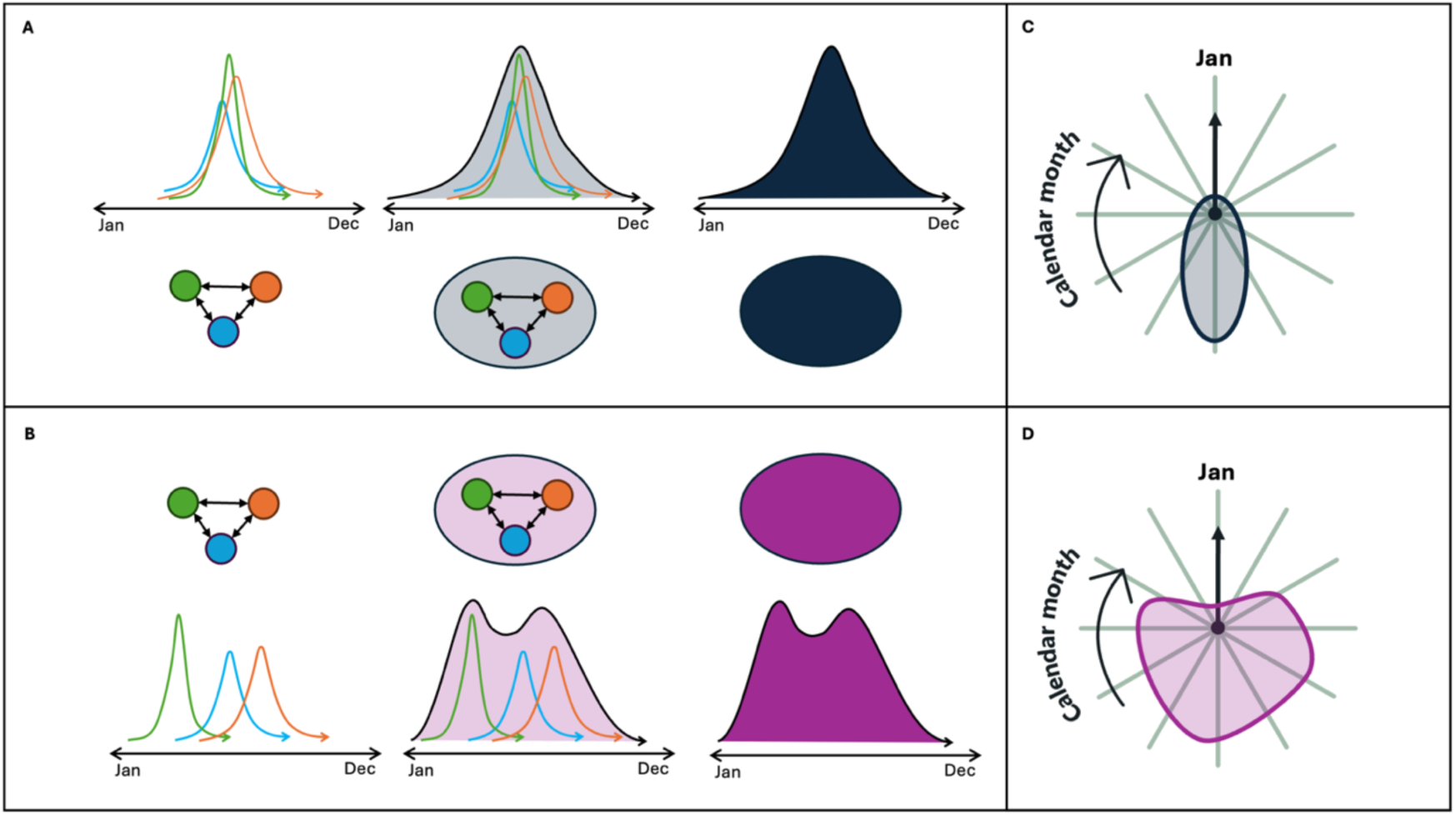
Representation of incidence time series and the resulting shape when plotted on a polar axis. Panel A shows the patch-level time series under a low vaccination coverage scenario in blue, green, and orange on the left, which are only observed at the metapopulation level (navy blue) on the right. Panel B shows the patch-level time series on the left, which are only observable at the metapopulation level (magenta) on the right. Panel C shows the resulting relative incidence from the observed data in a low coverage scenario on a polar axis, while Panel D shoes the resulting relative incidence from the observed data in a high coverage scenario on a polar axis.

We can leverage this expected broadening of the seasonal variation in incidence to indicate the transition from endemic circulation to metapopulation persistence in the overall dynamics. Here, we create a metric named roundness, defined as the area of the relative incidence plotted on a polar axis. We illustrate that roundness is positively correlated with vaccination coverage and the contribution of metapopulation extinction-colonization to local persistence. We first establish this through a stochastic metapopulation compartmental model. We then illustrate this phenomenon using historical measles surveillance data from the United States spanning the pre-vaccine era through disease elimination, and contemporary measles surveillance data in Nigeria, where there is substantial spatial variation in vaccination coverage, and subsequently, in measles transmission dynamics.

When a time series of incidence is plotted on a polar axis, such as in **Figure 1**, C and D, we can expect to see very different forms of the resulting plot under different transmission regimes. Under a scenario with persistent local transmission and little influence of metapopulation extinction/colonization dynamics, we expect to see a polygon with a smaller area and less roundness (**Figure 1**, C), while under a scenario where metapopulation rescue effects are influential, we expect to see a polygon with a rounder shape and a larger area (**Figure 1**, D).

## Data and Methods

### Roundness

We created vectors of scaled incidence (the maximum incidence for a given period was scaled to 1) for each year on a polar plane, where the angle represented the month or biweekly period, depending on data aggregation and availability, and the magnitude represented the relative number of cases compared to the maximum number of cases in that year. For example, the first biweekly period of the calendar year was represented as the cardinal direction North to 13.8° clockwise. The 14^th^ biweekly period, or the first biweekly period of the latter half of the calendar year, was represented as the cardinal direction South to 13.8° clockwise. In data available at a monthly level, the first monthly period was represented as the cardinal direction North to 30° clockwise, while the 7^th^ monthly period was represented as the cardinal direction South to 30° clockwise.

Using moving 3-year periods of scaled incidence vectors, we calculated the geometric mean of the minimum and maximum radii created by those vectors for each year. A larger geometric mean, or a higher value of roundness indicates that the maximum and minimum incidence values are similar, and that there is a reduced seasonal pattern of incidence. A smaller geometric mean, or a lower value of roundness, indicates that the maximum and minimum incidence values in a year are less similar, and that there is a more prominent seasonal patterns of incidence.

### Data

#### Simulation

We built a discrete-time, stochastic metapopulation compartmental (TSIR) model with 5 patches, adapting data used by *Dalziel et. al* on measles dynamics in the five boroughs of New York City in the pre-vaccine era (Bjørnstad et al., 2002; Dalziel et al., 2016). We further specified levels of vaccination coverage ranging from 0%-100% of new births for each run. We then compiled the numbers infected in each patch and in the aggregate across all patches at steady state (20 years).

In our TSIR model, the expected numbers of new infected individuals, λ, in *p* patches in generation t + 1 is defined as a vector as follows:

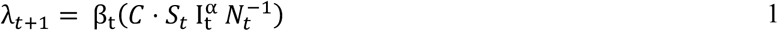

Where β_t_is the time-varying, seasonal transmission parameter adapted from Dalziel et. al. *C* is a matrix of dimensions *p* × *p* representing commuter-type coupling between patches. The values of *C* consist of a proportion of individuals, *c*, on the off-diagonals and 1 − *c* ∗ *p* on the diagonals. *S_t_* represents the vector of susceptibles in *p* patches at time *t*, 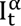 represents the vector of infected individuals in *p* patches at time *t*. The power *α* represents an adjustment made to correct for the use of a discrete-time model (Glass et al., 2003).

We then sample from a Poisson distribution with mean and variance λ*_p_*_,*t*+1_ to obtain the actual number of infected individuals *I_p_*_,*t*+1_ for time *t* + 1 and patch *p*.

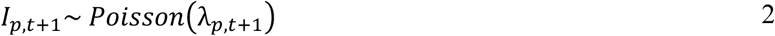

To ensure persistent transmission at the aggregate level, even if all patches go to 0, we introduced *Q* new cases from outside the metapopulation from a binomial draw as follows:

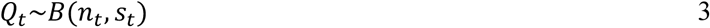

Where *n* is the size of the binomial draw and *s* is the probability of success at a 2-week infectious period *t*. For example, if *n_t_* = 1 and *s_t_* = 0.1, then there is a 10% probability of the introduction of 1 infected case to each patch in the metapopulation in that 2-week period. We constructed *Q_t_* in two ways: uniform and time-varying. In the uniform construction, the expected number of introduced cases for all 2-week periods, *t*, were identical, by setting all *n_t_* to be equal and all *s_t_* to be equal. In our simulations we used *n_t_* = 1 and *s_t_* = 0.07. A time-varying construction of *Q_t_* was developed to better reflect the realities of disease reporting and human mobility. If we consider our metapopulation as a jurisdiction (e.g., a country), population mobility from outside of the metapopulation may lead to introduced cases that may not be detected or reported by the jurisdiction of the metapopulation until further transmission has occurred. This can be especially true in areas with substantial cross-border mobility and reduced reporting capacity (WHO Ebola Response Team, 2014).

The number of susceptible individuals at time *t* + 1 is affected by births of susceptible individuals *B_t_*, the proportion of new births vaccinated *ρ*, and the infection process moving susceptible individuals to the infected class at time *t*.

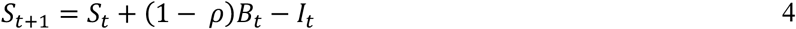

We varied *ρ* between 0 and 1 (corresponding to vaccination coverage at birth of 0% to 100%).

#### Historic measles time series

Monthly, state-wise counts of measles cases in the United States of America (US) were extracted from Project Tycho, ranging from years 1883 through 2001, covering the range of years including pre-vaccine endemic circulation, the introduction and adoption of the measles-containing vaccine, and measles elimination in the US (Orenstein et al., 2004; Van Panhuis et al., 2018). While we do not have comprehensive national or state-level estimates of vaccination coverage spanning this time period, coverage at the national level among 1-4 year old children increased from 40-60% in 1970 to above 90% by 1998 (Hutchins et al., 2004). Our data spans several major changes in the epidemiology of measles and its control. The MCV was introduced in 1963, and by 1966, the first elimination effort was introduced (Orenstein et al., 2004). While this and subsequent attempts to eliminate measles (1978 and 1993) were unsuccessful, these attempts dramatically reduced the incidence of measles, from 313 cases per 100,000 in 1956-1960, immediately prior to the introduction of the MCV, to 1.3 cases per 100,000. Later that decade, outbreaks in school-aged children led to a second dose being recommended for all children, which led to a further decline in incidence, before elimination was declared in 2000 (CDC, 2024).

#### Contemporary measles time series

Monthly, state-wise counts of suspected measles cases in Nigeria were extracted from aggregated data on healthcare-seeking visits, routinely submitted to the World Health Organization (WHO), encompassing the years 2007 through 2020. Suspected measles cases are defined as those with maculopapular rash (non-vesicular) and fever, and any of conjunctivitis, coryza, or cough (World Health Organization, 2018). Data were obtained from WHO measles surveillance database through a request to the Immunizations, Vaccines, and Biologicals (IVB) Department. These surveillance data are also publicly reported through routine measles situation reports published by the Nigeria Centre for Disease Control. However, the WHO database was used for this analysis because it allowed standardized extraction of historical data across the study period (Nigeria Centre for Disease Control and Prevention, 2025). Data on ADM1-level vaccination coverage in Nigeria were obtained from the Demographic and Health Surveys (DHS). Specifically, we used the percentage of children aged 12-23 months of age who received at least 1 dose of the measles containing vaccine (MCV) at any time in the past, according to a documented vaccination card or the child’s caregiver (Croft et al., 2018). As the DHS have historically been conducted approximately every 5 years, data for Nigeria were available for years 2013 and 2018.

### Calculation of metapopulation indicators

In addition to roundness, we calculated the following measures for the simulated and US time-series: incidence (also for Nigeria), proportion of time each patch was extinct (i.e., had 0 reported cases), proportion of patches 0 at any given time, maximum duration of extinction in any given year, and number of rescue events (i.e., a patch going from extinct at time *t* to extant at time *t* + 1).

## Results

### Simulation

Results from our simulation show that extinction and colonization dynamics tend to become more prevalent at higher levels of vaccination coverage upon running the model to equilibrium. Namely, the proportion of patches at 0 increases sharply as vaccination coverage increases, with values close to or at zero up to 92%. A sharp increase is observed above 95% coverage, where a majority of patches are extinct at any given time. The point at which this increase occurs varies at different levels of connectivity—at a higher degree of connectivity, we observe non-zero proportions of extinct patches at a higher level of coverage. Finally, the number of rescue events, or biweekly periods where a case was observed after a patch went extinct, increases dramatically as vaccination coverage increases. This phenomenon, too, varies by connectivity. at a higher degree of connectivity, we see a much steeper slope and a higher overall frequency of rescue events.

Changes in the values of roundness correspond with the increases in indicators of metapopulation dynamics. As coverage increases, we see an increase in roundness. Roundness increases from a base value of roughly 0.1 to a peak of roughly 0.3 as coverage increases (**Figure 2**, C). Higher levels of connectivity have a similar inflection point of coverage after which roundness rapidly increases, but show higher values of roundness at lower levels of coverage (**Figure 2**, B). Scaling the transmission parameter β so transmission is halved shows a similar pattern of relatively linear roundness values as coverage increases, followed by an inflection point and a rapid increase of roundness for increasing values of coverage. Scaling β so it is only 20% of the default value led to a relatively linear increase in roundness, indicating the presence of metapopulation dynamics even at low levels of coverage (**Error! Reference source not found.**). We also observe a smaller maximum slope and higher overall value of roundness in settings with lower transmission. Notably, roundness starts to increase rapidly at a level of vaccination coverage below a theoretical herd immunity threshold.

**Figure 2:**
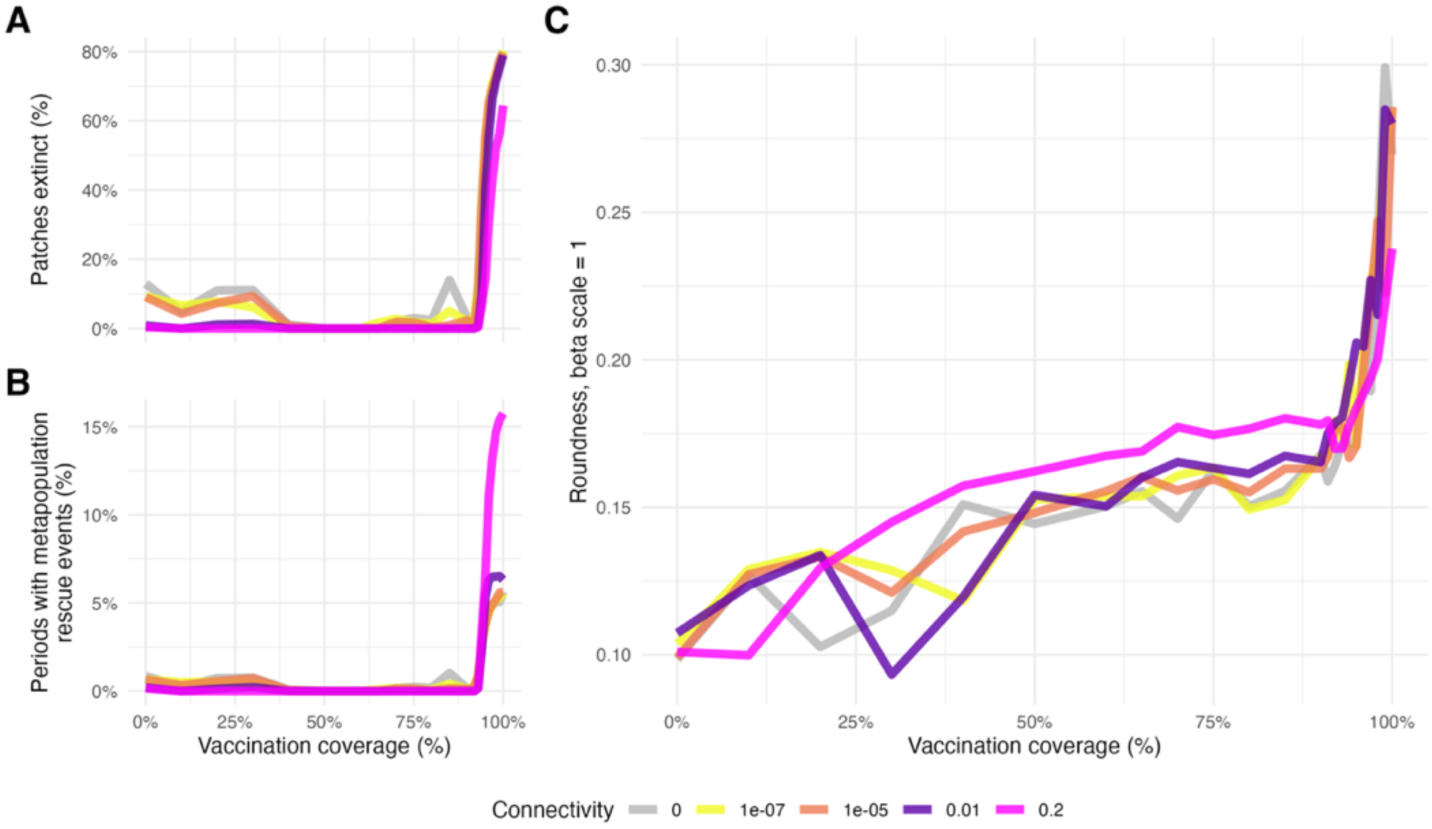
A: Proportion of patches at 0 by vaccination coverage. B: Frequency of metapopulation rescue events by vaccination coverage. C: roundness by vaccination coverage. Colored lines indicate model iterations with different between-patch connectivity.

### USA Time series

Measles incidence declined dramatically after the introduction of the vaccine, from multiple peaks of above 100,000 monthly cases, and an average of over 40,000 monthly cases in the 10 years prior to the introduction of the measles-containing vaccine in 1963, to under 10,000 monthly cases on average in the 10 years following the introduction of the vaccine.

Considering each state in the US as a patch in the metapopulation, the proportion of patches at 0 at any given month increased dramatically after the introduction of the vaccine, starting at 5% prior to the introduction of the vaccine and peaking at 100% in the 1990s (see **Figure 3**, A). Accordingly, the proportion of states experiencing metapopulation rescue (defined as monthly incidence increasing from 0 to above 0 in a given calendar year) also increased from ∼5% prior to the introduction of the vaccine to over 80% in the 1990s (**Figure 3**, B). Notably, roundness increased after the introduction of the vaccine, going from a value of ∼0.2 prior to the introduction of the vaccine to a value of ∼0.6 in the 1990s (**Figure 3**, C). We can also consider each state as a metapopulation itself. When we do so, we find that state-level roundness tracks roughly with national-level roundness in terms of magnitude, while leading the national-level roundness values by 1-2 years (**Figure 3**, C). Notably, we also see a continued increase in state-level roundness in the late 1980s onwards despite a drop in national-level roundness. This may be indicative of the resurgence of measles in the US in 1989 to 1991 in a handful of states, driving roundness down (as measured at the national level), while keeping roundness across most states high (Atkinson et al., 1992; Orenstein et al., 2004). We also see the effect of this resurgence in **Figure 3**, A, when the proportion of patches at 0 drops from over 75% before the resurgence to approximately 60% in some months after the resurgence.

**Figure 3:**
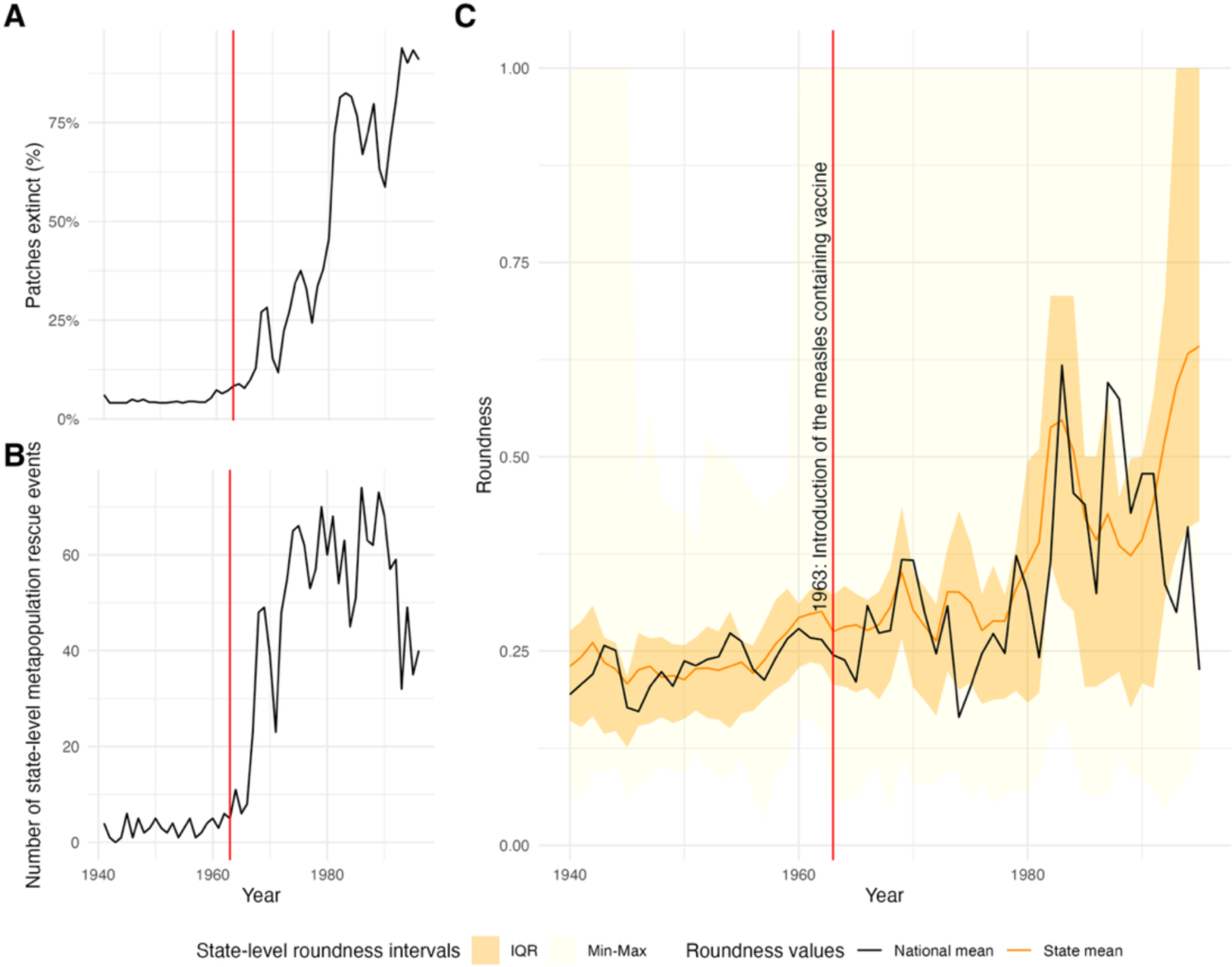
A: Proportion of states with 0 monthly cases by year. B: Number of metapopulation rescue events by year. C: Mean roundness at the US level by year as a solid black line, at the state level as a solid orange line, with the inter-quartile roundness shown in dark yellow and total range shown in light yellow. The red vertical line in all plots indicates 1963, the year that the measles containing vaccine was introduced in the US.

### Nigeria Time series

The goal of using the Nigeria time series was to show the variability in roundness across a spatial gradient of vaccination coverage, where we expected states within Nigeria with higher coverage to have correspondingly higher roundness values. State-level vaccination coverage in Nigeria increases along a north to south gradient (**Figure 4**, A). Considering each state as a separate metapopulation, we observe high variability in vaccination coverage, ranging from close to 0% to almost 90% at the state level. We also observe a trend of greater roundness corresponding to higher vaccination coverage (**Figure 4**, B). The spatial gradient of vaccination in Nigeria, where coverage tends to be higher in the southern states compared to the northern states, corresponds with the monthly distribution of relative incidence, with southern states showing visibly rounder within-year patterns of incidence compared to northern states (**Figure 4**, C).

**Figure 4:**
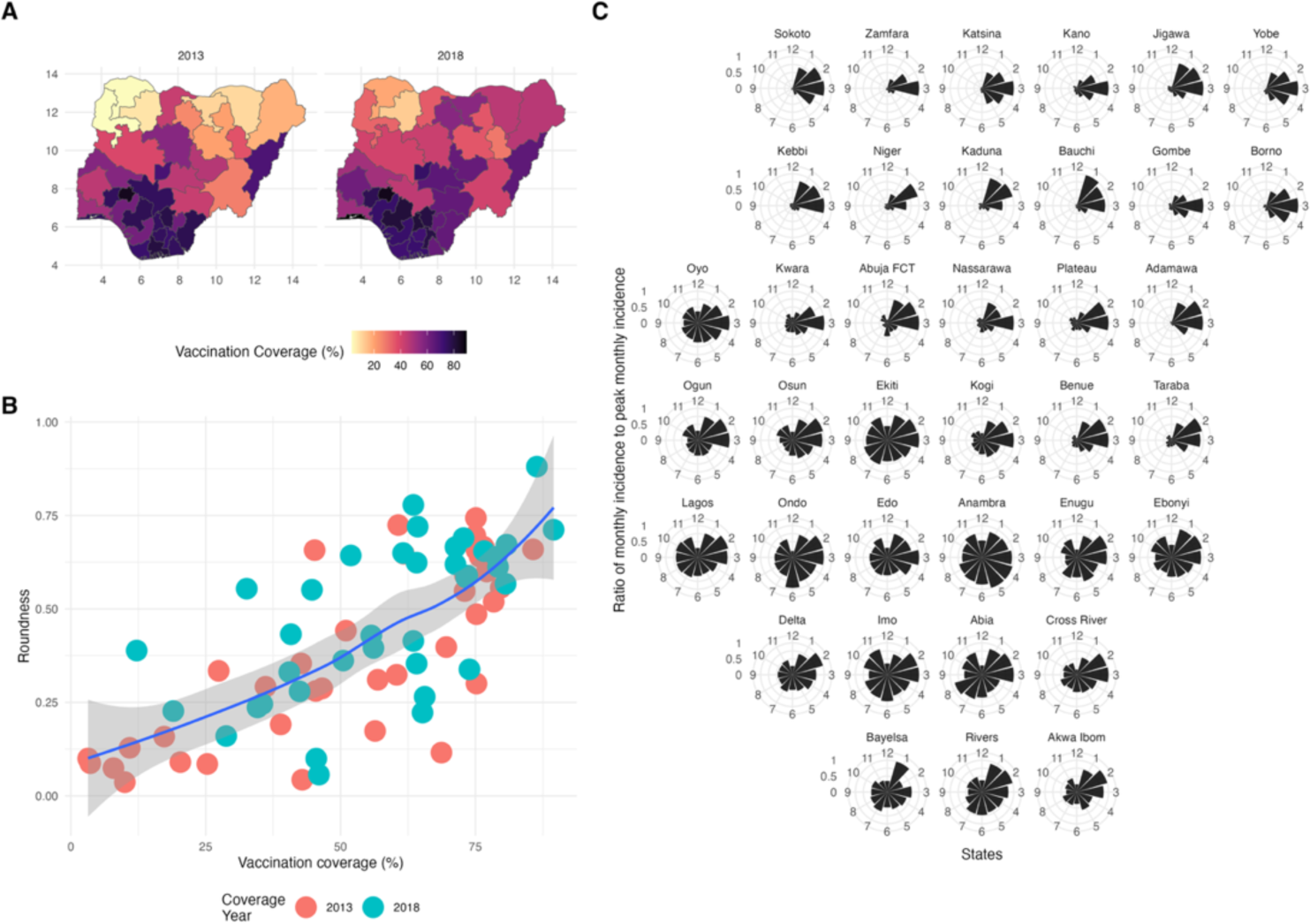
A: Vaccination coverage in Nigeria for years with available survey data—2013 and 2018. B: Correlation between state-level coverage and roundness for 2013 and 2018. C: Monthly incidence relative to peak incidence month for each state in Nigeria, arranged spatially.

## Discussion

We demonstrate that the changing seasonal distribution of disease incidence without an underlying change in the seasonality of transmission, captured by a novel measure called roundness, is indicative of the dominance of metapopulation extinction-colonization dynamics as vaccination coverage increases and incidence declines. Results from our simulations show that as vaccination coverage increases, patch-level indicators of metapopulation dynamics such as the proportion of patches extinct, and the number of rescue events per year, also increase. Similarly, roundness increases with increasing vaccination coverage, with a more gradual increase evident in lower transmission settings. We also note that roundness follows a trajectory that reflects between-patch dynamics. As coverage increases, roundness stays flat. At higher levels of coverage, metapopulation extinction-colonization dynamics start appearing, and it is at this point that we observe an “inflection point” in the value of roundness at roughly 94%, occurring at a level of coverage slightly below a theoretical herd immunity threshold for a similar mean value of the transmission parameter, *β* of 96.7%^1^. Roundness, once past an inflection point, appears to indicate the presence of metapopulation extinction-colonization dynamics. At very low levels of transmission (achieved by scaling all *β* values by 0.2), we see a more linear increase in roundness, indicating the presence of extinction-colonization dynamics even at low levels of vaccination coverage.

Using historical data from the United States, we see similar signatures of changing seasonality at the national level as observed in our simulations. This is further supported by state-level observations (e.g., increasing number of months with no cases, and increasing number of consecutive months with no cases) signaling a change in the dominant regime from endemic circulation toward metapopulation persistence after the introduction of the measles-containing vaccine. Our observations correspond with milestones in measles control in the USA, with sharp increases in roundness seen in 1966 and 1980, where elimination efforts were particularly strong (Orenstein et al., 2004). Notably, national-level roundness decreases in the late 1980s while state-level roundness continues to increase. This can be explained by severe measles outbreaks in largely-unvaccinated children of pre-school age in a few cities in the United States (Los Angeles, Houston, Milwaukee, Chicago, Dallas, and New York City) between 1989-1990, where there were over 40,000 cases reported in the two years, the highest since 1977 (Atkinson et al., 1992). These outbreaks would have disproportionately contributed to the burden of disease, and calculation of the roundness metric, skewing it downward at a national level. As most states were not affected by these outbreaks, state-level roundness remained high.

Within states in Nigeria, where metapopulation dynamics cannot be directly observed, we see a strong correlation between roundness and coverage. While we cannot rule out the effect of changing seasonality over time in our observed time series data, it is striking that the patterns that we do see are consistent with known gradient and vaccination control over time and space and with each other.

This study is situated in work that aims to understand the level of control required where there is no longer any persistence of the pathogen in the metapopulation (i.e., elimination of the pathogen), akin to a herd immunity threshold in a single population. This threshold can depend on the size of the metapopulation and its constituent patches, vaccination coverage, connectivity between patches, and spatial and temporal heterogeneity between patches (Kortessis et al., 2025). Other approaches that have aimed to study this phenomenon include critical slowing down theory, canonical path, and the early warning system (EWS) indicators such as autocorrelation (Brett et al., 2020; Graham et al., 2019; O’Regan & Drake, 2013). Somewhat similarly to roundness, these indicators also rely on changing properties of incidence values over time to detect changes in the dynamics of the system from steady state to either emergence or elimination. In addition to signaling shifts in the regime, our measure of roundness also indicates how the regime is changing—a system with endemic transmission has much lower, and much more stable values of roundness compared to a system propped up by metapopulation persistence and extinction/colonization events.

An advantage of roundness is that it can be measured even when case reporting is imperfect. We tested under-reporting by artificially reducing case counts from our simulations by 60% and found no qualitative difference in roundness values created from the artificially under-reported data (Appendix B). Even if a low proportion of cases are seen and subsequently reported, roundness can be used to assess whether a transition away from endemic transmission is occurring. The use of roundness can also be supplemented by the use of other measures that are demonstrably linked to immunity, such as the mean age of cases, the proportion of cases testing positive for ongoing infection, and the vaccination history of cases when case counts are low, to assess whether incidence is truly low and a transition to metapopulation persistence and elimination is occurring (Bhatia et al., 2025).

One limitation of this study is the roundness may not be interpretable on an absolute scale, as the shape and strength of seasonal drivers affecting transmission can vary between places. For example, term time forcing and agricultural cycles, factors that affect the seasonality of transmission, vary between places (Bharti et al., 2011; Ferrari et al., 2010). While we cannot exclude the probability that differences are due to changes in underlying seasonality over time, the relationship between roundness and control efforts holds in a range of different seasonal transmission regimes (USA through the 20^th^ century vs. individual states in Nigeria in the 21^st^ century). If there is data collection on incidence prior to, during, and after the introduction of large-scale vaccination programs, then the roundness value can be well characterized for a given place, such as was done in the US. The key utility of this measure is to allow comparison in a metapopulation over time, or to allow comparison of different metapopulations in a larger system.

## Conclusion

We show that a novel measure, roundness, can be used to effectively track regional transitions from endemic circulation to metapopulation persistence without using sub-regional data on immunity, which may not be available or reliable. We first show how roundness changes in a connected metapopulation with varying levels of vaccination coverage using simulations. We then consider the entirety of the United States as one metapopulation and show that roundness, as well as indicators suggesting increases in metapopulation-based dynamics both increased over time after the introduction of the measles-containing vaccine until elimination was achieved. Finally, we consider each state in Nigeria as a metapopulation and show that roundness at the state level is highly correlated with vaccination coverage, further suggesting the difference in dynamics between high-coverage, high-roundness states and lower-coverage low-roundness states.

Understanding how disease systems change over time is important in managing those systems toward elimination and extinction. However, relying only on reported cases can be misleading in situations where the ability to report cases consistently changes, such as resource limitations or conflict. It is in situations like these where populations can be more vulnerable due to a reduced capacity to vaccinate against, report, and manage vaccine-preventable diseases such as measles. Roundness is a novel method to assess how systems are changing, and the use of roundness can offer insights not only on how disease systems are changing, but *where* they are changing, and where efforts may need to be directed.

## Data Availability

Nigeria measles incidence data can be downloaded at https://ncdc.gov.ng/data or through a request to the Immunizations, Vaccines, and Biologicals (IVB) Department at the World Health Organization (WHO).

https://github.com/objornstad/epimdr2

https://www.tycho.pitt.edu/dataset/api/

https://ncdc.gov.ng/data

## Footnotes

1 Due to the seasonality in transmission inherent in our model, however, we can only calculate a theoretical herd immunity threshold based on the mean *β* of 30.05, leading to a herd immunity threshold of 96.7%. A high amplitude of seasonality of *β* may lead to continued local or global extinction events, making a direct comparison difficult.

