## Supplementary Materials for "Characterizing the relative strength of metapopulation dynamics in measles persistence"

**Supplementary Materials for *Characterizing the relative strength of metapopulation dynamics in measles persistence* (Bhatia, McKee, and Ferrari)**

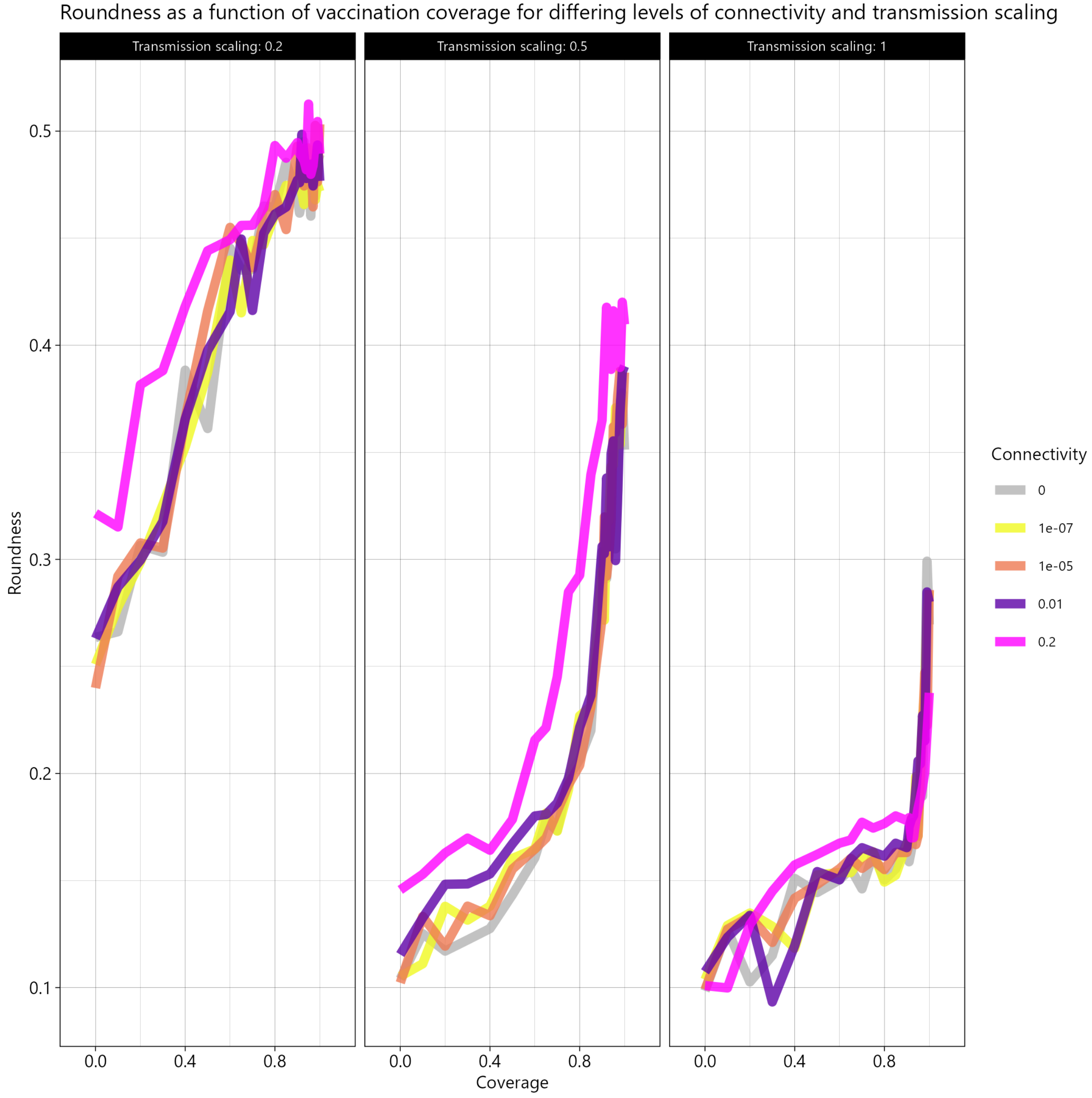


*Supplementary Figure 1: Roundness (y-axis) at different levels of coverage (x-axis), connectivity among patches (colors) and scaling of the transmission parameter, beta (panels).*

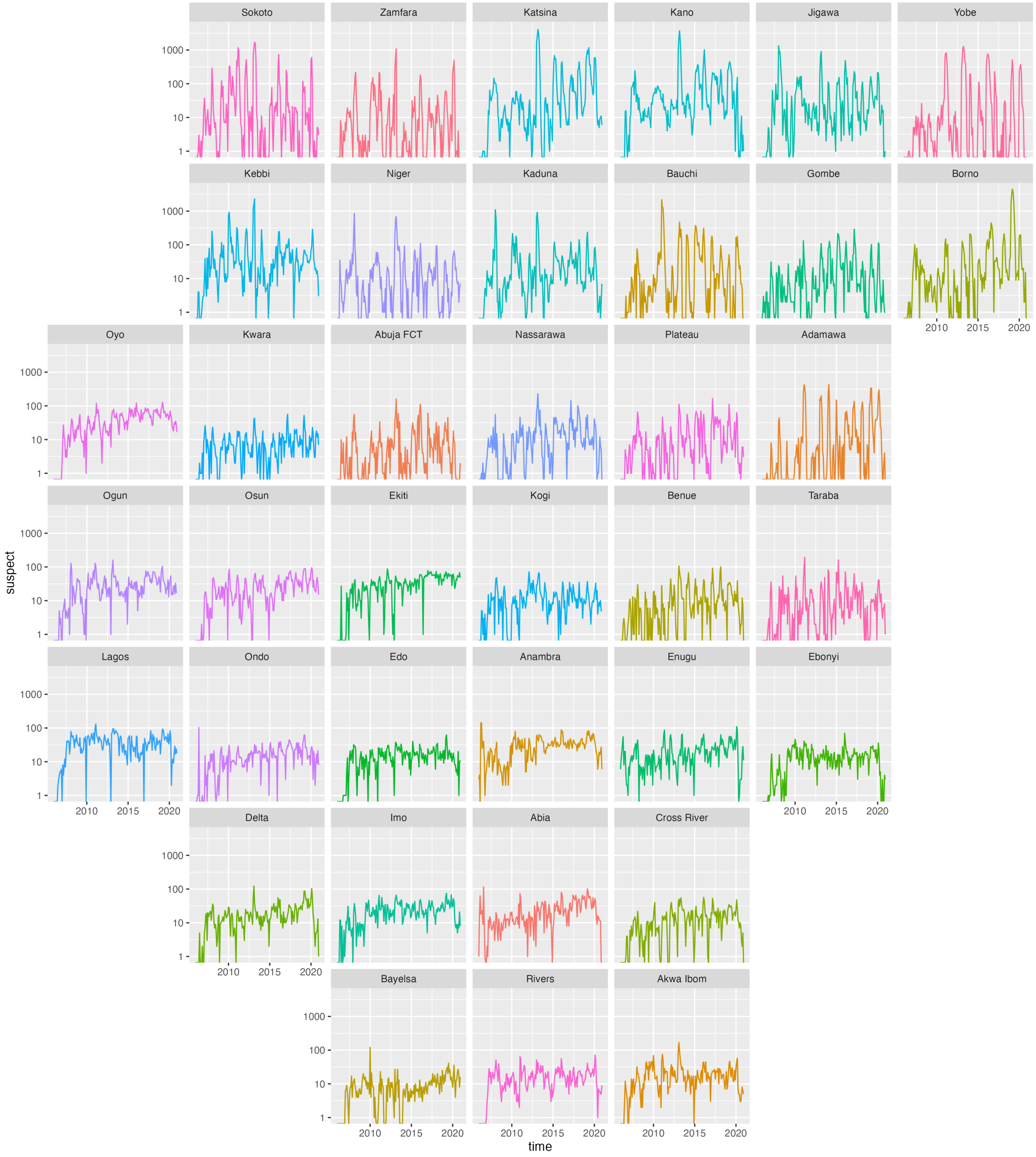


*Supplementary Figure 2: Suspect cases of measles in Nigeria by month, log-scale.*

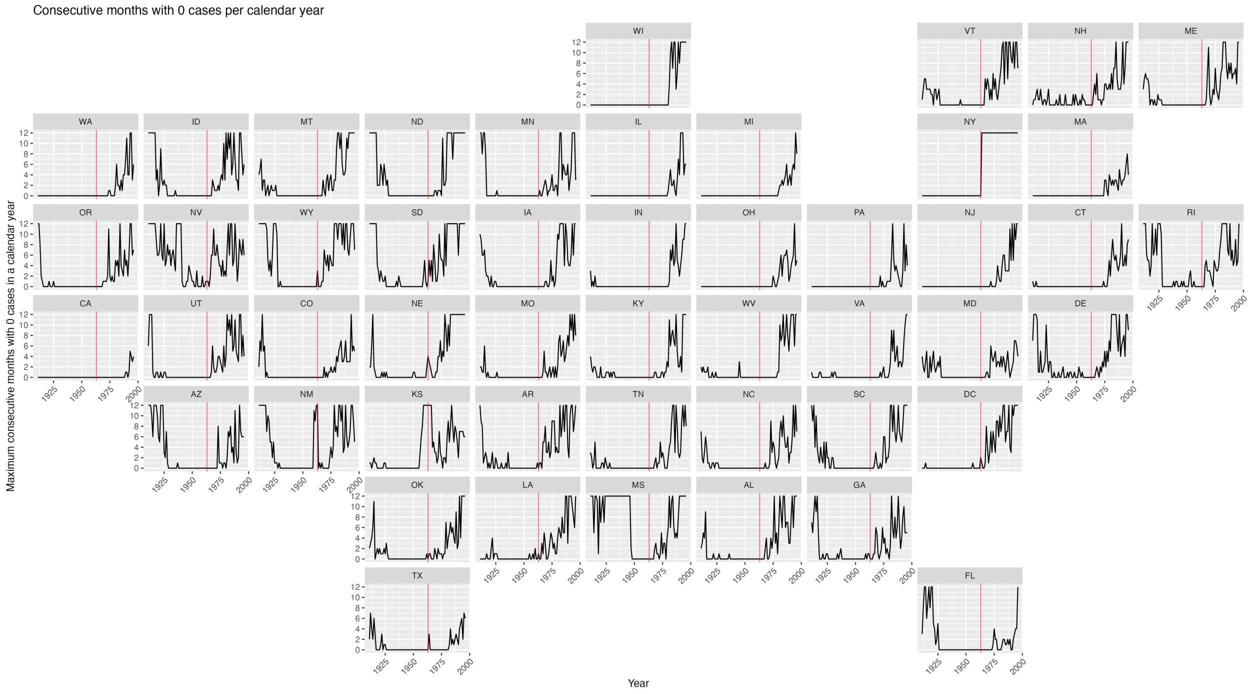


*Supplementary Figure 3: Consecutive months with 0 cases of measles per calendar year by state. The vertical red line at 1963 indicates the introduction of the measles containing vaccine in the US.*

### Testing for robustness with under-reporting

We tested for under-reporting by drawing from a binomial distribution 100 times with a probability of success of 40%. This produced a value from 0-100. We multiplied this value by 0.01 to scale it between 0 and 1, and then multiplied the simulated incidence by this value, thus scaling it.
